# Bayesian forecasting for intravenous tobramycin dosing in adults with Cystic Fibrosis using one versus two serum concentrations in a dosing interval

**DOI:** 10.1101/2020.12.31.20249095

**Authors:** Philip G. Drennan, Yann Thoma, Lucinda Barry, Johan Matthey, Sheila Sivam, Sebastiaan J. van Hal

**Affiliations:** Department of Microbiology and Infectious Diseases, Royal Prince Alfred Hospital, Sydney, Australia; School of Management and Engineering Vaud (HEIG-VD), University of Applied Science Western Switzerland (HES-SO), Yverdon-les-Bains, Switzerland; Department of Respiratory Medicine, Royal Prince Alfred Hospital, Sydney, Australia; University of Sydney, Australia

**Author notes:** Corresponding author: Philip Drennan, email address for correspondence.

## Abstract

**Background:** Intravenous tobramycin requires therapeutic drug monitoring (TDM) to ensure safety and efficacy when used for prolonged treatment, as in infective exacerbations of Cystic Fibrosis (CF). The 24 hour area under the concentration time curve (AUC_24_) is widely used to guide dosing, however there remains variability in practice around methods for its estimation.

**Objectives:** To determine the potential for a sparse sampling strategy using a single post-infusion tobramycin concentration and Bayesian forecasting, to assess the AUC_24_ in routine practice.

**Methods:** Adults with CF receiving once daily tobramycin had paired concentrations measured 2 hours (c_1_) and 6 hours (c_2_) following end of infusion as routine monitoring. We estimated AUC_24_ exposures using Tucuxi, a Bayesian forecasting application incorporating a validated population pharmacokinetic model. We performed simulations to estimate AUC_24_ using the full dataset using c_1_ and c_2_, compared to estimates using depleted datasets (c_1_ or c_2_ only), with and without concentration data from earlier in the course. We assessed agreement between each simulation condition and the reference graphically, and numerically using median difference (Δ) AUC_24_, and (relative) root mean square error (rRMSE) as measures of bias and accuracy respectively.

**Results:** 55 patients contributed 512 concentrations from 95 tobramycin courses and 256 TDM episodes. Single concentration methods performed well, with median ΔAUC_24_ <2 mg.h.l^-1^ and rRMSE of <15% for sequential c_1_ and c_2_ conditions.

**Conclusions:** Bayesian forecasting, using single post-infusion concentrations taken 2-6 hours following tobramycin administration can adequately estimate true exposure in this patient group and are suitable for routine TDM practice.

**Key Points:**

- In stable adult patients with Cystic fibrosis without significant renal impairment, Bayesian forecasting allows accurate estimation of tobramycin AUC_24_ using a single blood sample taken 2-6 hours post-infusion with acceptable accuracy, especially when including prior measured concentrations.
- A single sample approach with Bayesian forecasting is logistically less complicated than a two-sample approach, and could facilitate best-practice TDM in the outpatient setting.
- A more intensive sampling strategy with Bayesian forecasting using two tobramycin concentrations in a dosing interval should be considered in unstable patients, or where observed concentrations deviate significantly from model predictions.

## 1. Introduction

Life expectancy for patients with Cystic Fibrosis (CF) continues to improve, owing to a range of strategies to maintain lung function, including aggressive treatment of infective pulmonary exacerbations due to *Pseudomonas aeruginosa*.(1) Intravenous tobramycin is commonly prescribed for this purpose.(2) Well-known toxicities of tobramycin include nephrotoxicity and ototoxicity, and there is increasing interest in minimising these long-term sequelae, which are at least partly attributable to cumulative lifetime exposure.(2–4) Once-daily tobramycin dosing is increasingly becoming standard practice in many CF centres, however centres differ with regard to therapeutic drug monitoring (TDM) practices, which range from simple trough concentration and nomogram methods, to more direct monitoring of the 24 hour area under the concentration time curve (AUC_24_).(5–7) At our centre (Royal Prince Alfred Hospital, Sydney, Australia), we perform AUC_24_ based monitoring (target AUC_24_ 100 mg.h.l^-1^) using a log-linear regression (LLR) method, which requires two-concentrations within a dosing interval and simple pharmacokinetic calculations coded into a Microsoft Excel spreadsheet.(8) Although simple and accurate, significant attention is required to coordinate correct application of the TDM protocol, including obtaining two blood samples at the appropriate times, and ensuring the dosing and monitoring information is recorded accurately.(9,10) Thereafter, clinicians performing the AUC_24_ estimation must perform the required calculations in a timely manner in order to return dosing advice prior to the next dose. There is increasing interest in the ambulatory management of pulmonary exacerbations, however it is difficult and impractical to collect two post-infusion blood samples in the outpatient setting. A simple, accurate method of AUC_24_ estimation to inform ongoing dosing using a single blood sample may therefore greatly increase the efficiency of this TDM practice, broaden its utility to outpatient settings, and improve patient acceptability.

Bayesian forecasting, which incorporates information from population pharmacokinetic models with one or two measured post-infusion concentrations has been shown to accurately characterise tobramycin pharmacokinetics in adults with CF, compared to results derived from intensive sampling.(11,12) In a small study of 12 adults receiving once-daily tobramycin, Bayesian methods resulted in less biased and more precise AUC_24_ estimation than a LLR method, and one- and two-sample Bayesian strategies were similarly accurate.(11) The authors of this study considered all three methods potentially suitable for routine TDM. The aim of the current study was to examine the potential generalisability of this finding by studying a larger group of adult patients, representative of those typically managed in CF centres. We sought to determine the agreement in AUC_24_ estimation between a one-sample and a two-sample approach using Bayesian forecasting, in order to determine the potential suitability of a one-sample Bayesian forecasting approach for routine TDM in this group of patients.

## 2. Methods

### 2.1. Patients and data

We collected data retrospectively from adults (>18 years) with CF who received once-daily intravenous tobramycin for pulmonary exacerbations, as part of routine practice between January and December 2018. Clinical staff were instructed to take blood samples for the measurement of tobramycin concentrations at two time points (hereafter designated t_1_ and t_2_) at approximately 2 hours (c_1_) and 6-8 hours (c_2_) following the end of the infusion (0.5 hours). Dosing and drug concentration information, in addition to a patient’s height, weight, and the most recent creatinine measurement, was submitted to the TDM monitoring service for estimation of pharmacokinetic parameters and subsequent dosing advice, in order to achieve a target AUC_24_ of 100mg/L (acceptable range 80-120mg.h.l^-1^). In routine practice our TDM service currently uses a log linear regression method for AUC_24_ estimation, with proportional dose adjustment for subsequent dose recommendations.(8)

We recruited all patients with CF contributing at least one pair of tobramycin concentrations, which constituted a single TDM episode. We excluded episodes where c_2_ was below the lower limit of quantification, and where any of the concentration measurements was considered unreliable based on clinician assessment at the time of the TDM episode—for example where c_1_ was higher than c_2_. These cases were identified by the clinician performing the TDM, and prompted a request to repeat the measurements the following day.

The study was approved by the Sydney Local Health District Ethics Committee (ref X19-0168) with a waiver of informed consent due to retrospective collection of routinely collected data.

### 2.2. Tobramycin assay

Plasma tobramycin concentrations were determined using immune spectrophotometric assay on a Roche/Hitachi cobas c system platform with LLOQ of 0.33 mg l^-1^ and intra- and inter-day assay coefficients of variation <10%.

### 2.3. Simulation of sampling regimens

We simulated the potential effect of different sparse-sampling strategies on PK parameter and AUC_24_ estimates using the Tucuxi Bayesian forecasting software (www.tucuxi.ch), using a previously validated population pharmacokinetic model for adults with CF by Hennig et al.(12–14) Patient age, height, weight, and serum creatinine were covariates for the underlying population pharmacokinetic model used to generate the *a priori* pharmacokinetic parameter estimates. We used the full set of sequential concentration pairs (S_c1c2_) available at the time of each TDM episode as the reference (‘REF’), and compared this to one of five simulated sampling conditions (‘SIM’) which incorporated less information:

a. c_1_ and c_2_ without sequential prior concentration measurements from the same TDM episode (*NS*_*c1c2*_),
b. c_1_ measurements only, using sequential information from previous c_1_ measurements (*S*_*c1*_); c) c_1_ measurements only, without prior sequential measurements (NS_c1_); d) c_2_ measurements only, with sequential measurements (S_c2_; e) c_2_ measurements only, without sequential measurements (NS_c2_). We used customised scripts to perform the simulations using the Tucuxi engine, to reduce the risk of errors and to ensure reproducibility of the results.

### 2.4. Statistical analysis and measures of agreement

We performed statistical analysis using R (version 3.3.2, R Foundation for Statistical Computing, Vienna, Austria) implemented in the RStudio environment (version 1.0.136, RStudio Team (2016), RStudio, Inc., Boston, MA). As a measure of bias we estimated agreement of the PK parameters, and AUC_24_ between the reference and each of the simulated conditions for each TDM episode. The main parameter of interest was the estimated AUC_24_, calculated from the estimated clearance and the administered dose according to equation (1):

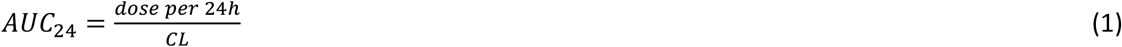

We calculated absolute differences in AUC_24_ estimates between each of the simulation conditions and the reference method according to equation (2):

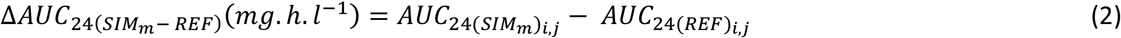

The relative difference in exposure estimates was calculated as per equation (3):

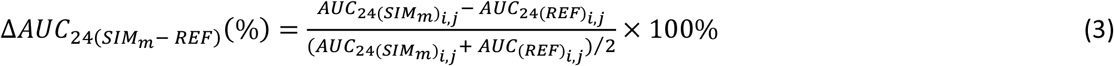

Where 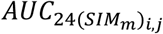 denotes the estimated AUC_24_ calculated using data from simulation m (*NS*_*c1c2*_, NS_c1_, S_c1_, NS_c2_, S_c2_) for the *i*th individual on the *j*th TDM episode, and 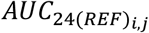 denotes the estimated AUC_24_ calculated using the all prior concentration data available for individual *i* at the time of the *j*th TDM episode. For the purposes of this study separate admissions were treated as independent cases.

As a measure of accuracy of the simulated method with the reference we calculated the root mean square error (RMSE, *mg. h. l*^−1^) and the relative RMSE (rRMSE, %) according to equations (4), and (5):

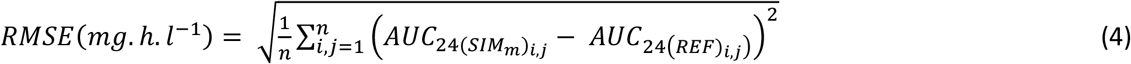

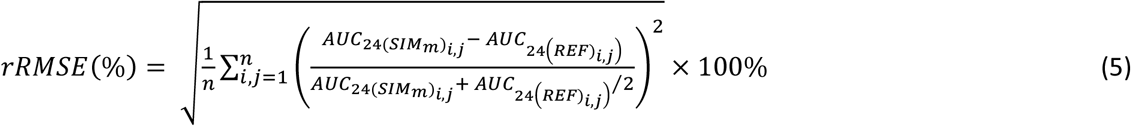

Where n is the total number of TDM episodes. We explored agreement of simulated conditions with the reference overall, and according to number of previous TDM episodes graphically, using boxplots and relative difference plots.(15)

## Results

### 2.5. Patients and data

Between January 1st and December 21^st^ 2018, 54 patients were admitted on 95 occasions for an infective exacerbation of their CF and received a course of tobramycin as part of their antimicrobial therapy. There were 256 TDM episodes (range 1-5 per course) and 512 concentrations. The median age was 31 years (IQR 22-40.25), and 23 (43%) patients were male. Tobramycin was given for a median 13 days (IQR 11-14). Demographics are summarised in table 1. Dosing and monitoring data, and pharmacokinetic parameter estimates for each TDM episode are shown in table 2 and figure 1. The median dose of tobramycin was 8.6 mg.kg^-1^ (IQR 7.1-10). Within a dosing interval, c_1_ was measured at a median of 2 h (IQR 2 – 2.6), and c_2_ at a median of 6.2 h (IQR 6-6.7). Across all TDM episodes the median estimated AUC_24_ using the Bayesian forecasting approach with all available data was 94.2 mg.h.L^-1^ (IQR 75.3-112.9).

**Table 1:** Baseline demographics and admissions

|  |  |
| --- | --- |
| Patients | 54 |
| Age (years), median (IQR) | 31 (22, 40.3) |
| Male, n (%) | 23 (42.6) |
| Height (cm), median (IQR) | 166 (160, 171) |
| Weight (kg), median (IQR) | 56 (50, 65) |
| Admissions/patient, median (range) | 1 (1, 6) |
| Admissions, total | 95 |
| Creatinine on admission (umol/L), median (IQR) | 64 (53, 77) |
| CRP on admission (mg/L), median (IQR) | 28 (7.9, 64.8) |
| T>38C on admission, n (%) | 13 (14.1) |
| Length of tobramycin course (days), median (IQR) | 13 (11, 14) |

**Table 2:** Tobramycin concentrations and parameter estimates for each TDM episode (n=256)

| Measurement/parameter estimate | Median (IQR) |  |
| --- | --- | --- |
| Dose administered (mg) | 520.0 | (400, 560) |
| Dose per kg (mg.kg<sup>-1</sup>) | 8.6 | (7.1, 10) |
| t<sub>1</sub> (h) | 2.0 | (2, 2.2) |
| t<sub>2</sub> (h) | 6.2 | (6, 6.7) |
| c<sub>1</sub> (mg.L<sup>-1</sup>) | 11.5 | (9.3, 14) |
| c<sub>2</sub> (mg.L<sup>-1</sup>) | 2.9 | (2.2, 3.9) |
| V<sub>1</sub> (L.70kg<sup>-1</sup>) | 18.4 | (15, 21.5) |
| V<sub>2</sub> (L.70kg<sup>-1</sup>) | 8.9 | (7.9, 10) |
| Q (L.h<sup>-1</sup>.70kg<sup>-1</sup>) | 1.5 | (1.3, 1.8) |
| CL (L.h<sup>-1</sup>.70kg<sup>-1</sup>) | 6.2 | (5.4, 7.5) |
| AUC<sub>24</sub> (mg.h.l<sup>-1</sup>) | 94.2 | (75.3, 112.9) |

**Figure 1:**
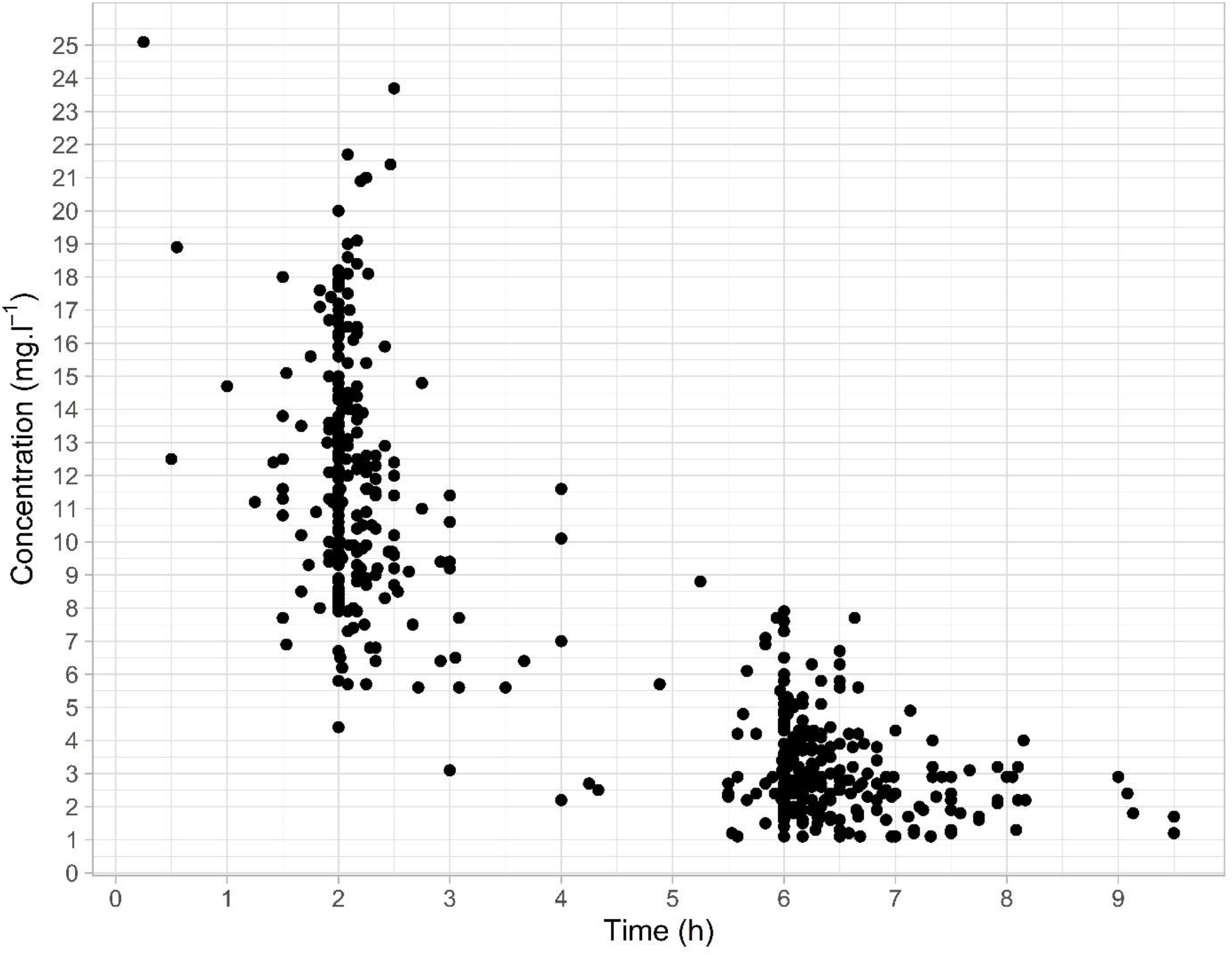
Time versus tobramycin concentration (n=512)

### 2.6. Comparison of simulation methods

Figure 2 shows the relative difference plot of ΔAUC_24_ (%) for each of the simulation conditions. There was no clear trend in agreement according to estimated AUC_24_. Table 3 and figure 3 show the estimated tobramycin exposure for the reference condition based on the full data set (Sc_1_c_2_) and each of the simulation conditions, and differences between the methods, pooled across all TDM episodes (n=256). AUC_24_ estimates were on average modestly greater than the reference condition, with median ΔAUC_24_ <5% compared to Sc_1_c_2_ for all conditions except NS_c1_. rRMSE of all methods was <15% of the reference AUC_24_. Large differences in estimated exposure, defined arbitrarily as ΔAUC_24_>30% were rare in all conditions but occurred least frequently in the S_c1_ condition (1.6% of episodes). Table 4 and figure 4 show comparisons of NS_c1c2_, S_c1_, and S_c2_ with the reference condition, stratified by TDM episode. As expected for NS_c1c2_, both ΔAUC_24_ and rRMSE increased for subsequent TDM episodes, as this condition only considered the most recent available data. For S_c1_ and S_c2_, rRMSE improved modestly for second and subsequent TDM episodes, and was <15% for all TDM episodes

**Table 3:** Comparison of simulation conditions

| Simulation condition | AUC<sub>24</sub> (mg.h.l<sup>-1</sup>) |  | ΔAUC<sub>24</sub> (mg.h.l<sup>-1</sup>) |  | ΔAUC<sub>24</sub> (%) |  | ΔAUC<sub>24</sub>>30% | RMSE (mg.h.l<sup>-1</sup>) | rRMSE (%) |
| --- | --- | --- | --- | --- | --- | --- | --- | --- | --- |
| Sc1c2 | 94.2 | (75.3, 112.9) | (ref) |  |  |  |  |  |  |
| NSc1c2 | 95.9 | (76.6, 118.1) | 0.0 | (0, 7.2) | 0.0 | (0, 7.6) | 8 (3.2) | 12.3 | 10.9 |
| Sc1 | 96.8 | (79.4, 112.5) | 1.6 | (-3.6, 7.1) | 1.7 | (-3.3, 8.4) | 4 (1.6) | 11.3 | 11.1 |
| NSc1 | 97.8 | (79.2, 119.8) | 4.9 | (-0.6, 13) | 5.3 | (-0.7, 13.6) | 9 (3.6) | 16.7 | 14.2 |
| Sc2 | 95.5 | (75.2, 116.8) | 1.2 | (-3.7, 6.7) | 1.8 | (-4.4, 6.8) | 7 (2.8) | 13.1 | 11.4 |
| NSc2 | 96.0 | (74.7, 119.1) | 3.1 | (-3.7, 9.5) | 3.5 | (-4.5, 10.5) | 7 (2.8) | 15.5 | 13.0 |

Data are expressed as median (IQR) or n (%)

**Table 4:** Comparison of simulation conditions according to number of preceding TDM episodes

| Simulation condition | TDM episode | $\Delta AUC_{24}$ (mg.h.l<sup>-1</sup>) | | $\Delta AUC_{24}(\%)$ | | RMSE (mg.h.l<sup>-1</sup>) | rRMSE (%) |
| --- | --- | --- | --- | --- | --- | --- | --- |
| NSc1c2 | first | 0.0 | (0, 0) | 0.0 | (0, 0) | 0.0 | 0.0 |
|  | second | 2.7 | (-3.2, 7.8) | 3.3 | (-3.2, 8.7) | 12.2 | 11.7 |
|  | third or greater | 8.1 | (-0.5, 18.4) | 8.3 | (-0.5, 14.8) | 19.1 | 16.1 |
| Sc1 | first | 3.1 | (-1.6, 10.2) | 5.0 | (-2, 12.1) | 12.8 | 13.0 |
|  | second | 1.3 | (-4.1, 5.6) | 1.4 | (-3.5, 6) | 10.8 | 9.9 |
|  | third or greater | -0.7 | (-5.6, 4.3) | -0.7 | (-5.9, 3.8) | 9.6 | 9.4 |
| Sc2 | first | 0.2 | (-6.2, 4.3) | 0.6 | (-7.3, 5.8) | 11.8 | 11.6 |
|  | second | 1.1 | (-2.7, 8.2) | 1.9 | (-2.8, 7.7) | 14.7 | 11.5 |
|  | third or greater | 2.3 | (-1.4, 9.3) | 2.5 | (-1.3, 10) | 12.8 | 11.1 |

Data are expressed as median (IQR)

**Figure 2:**
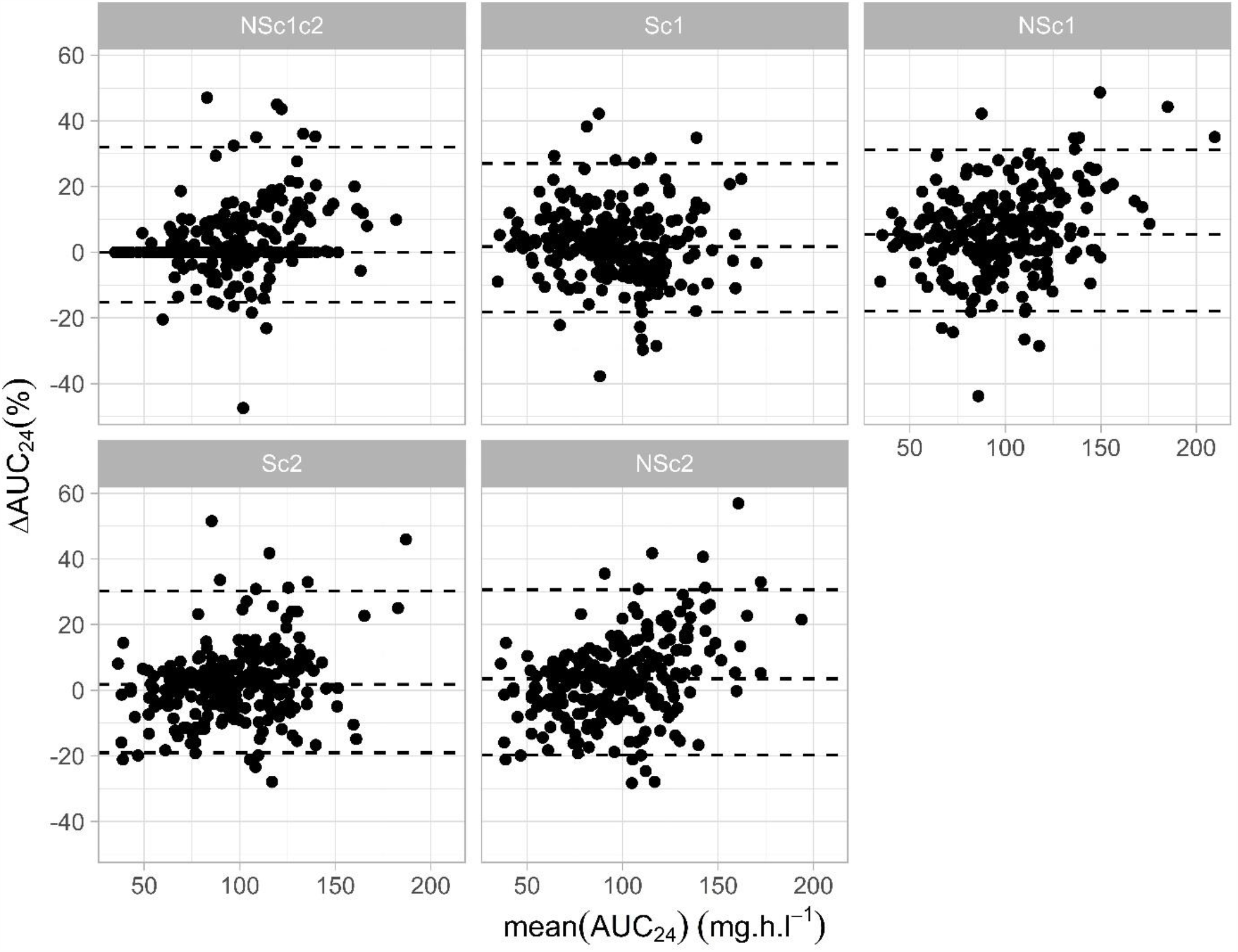
Relative difference plot of mean estimated AUC_24_ versus difference in AUC_24_ (%) for the reference method (full dataset) versus each simulated condition, with median difference and 2.5^th^ and 97.5^th^ percentiles (dashed lines)

**Figure 3:**
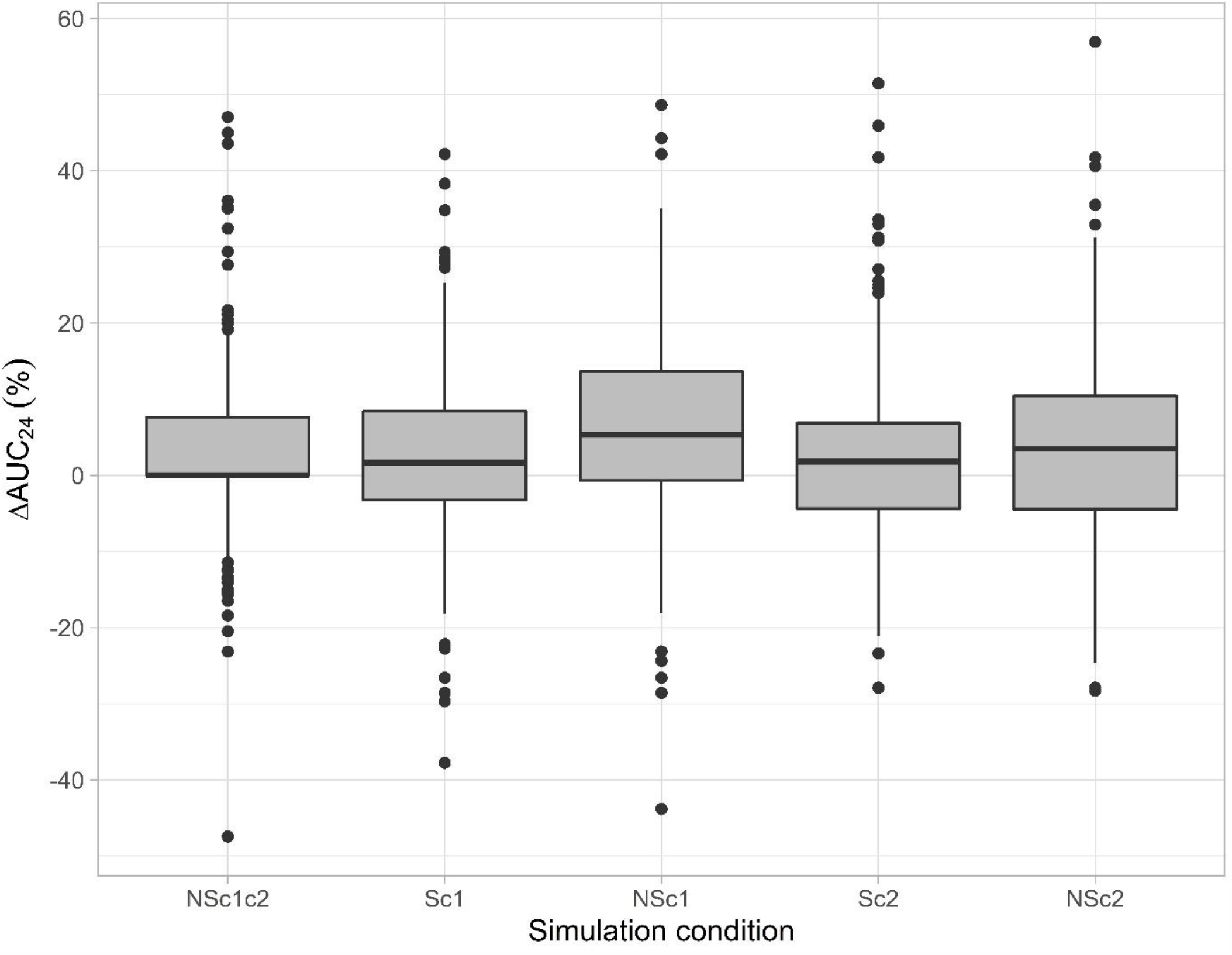
Distribution of differences in estimated AUC_24_ (%) for full data set (reference condition) versus each simulation condition

**Figure 4:**
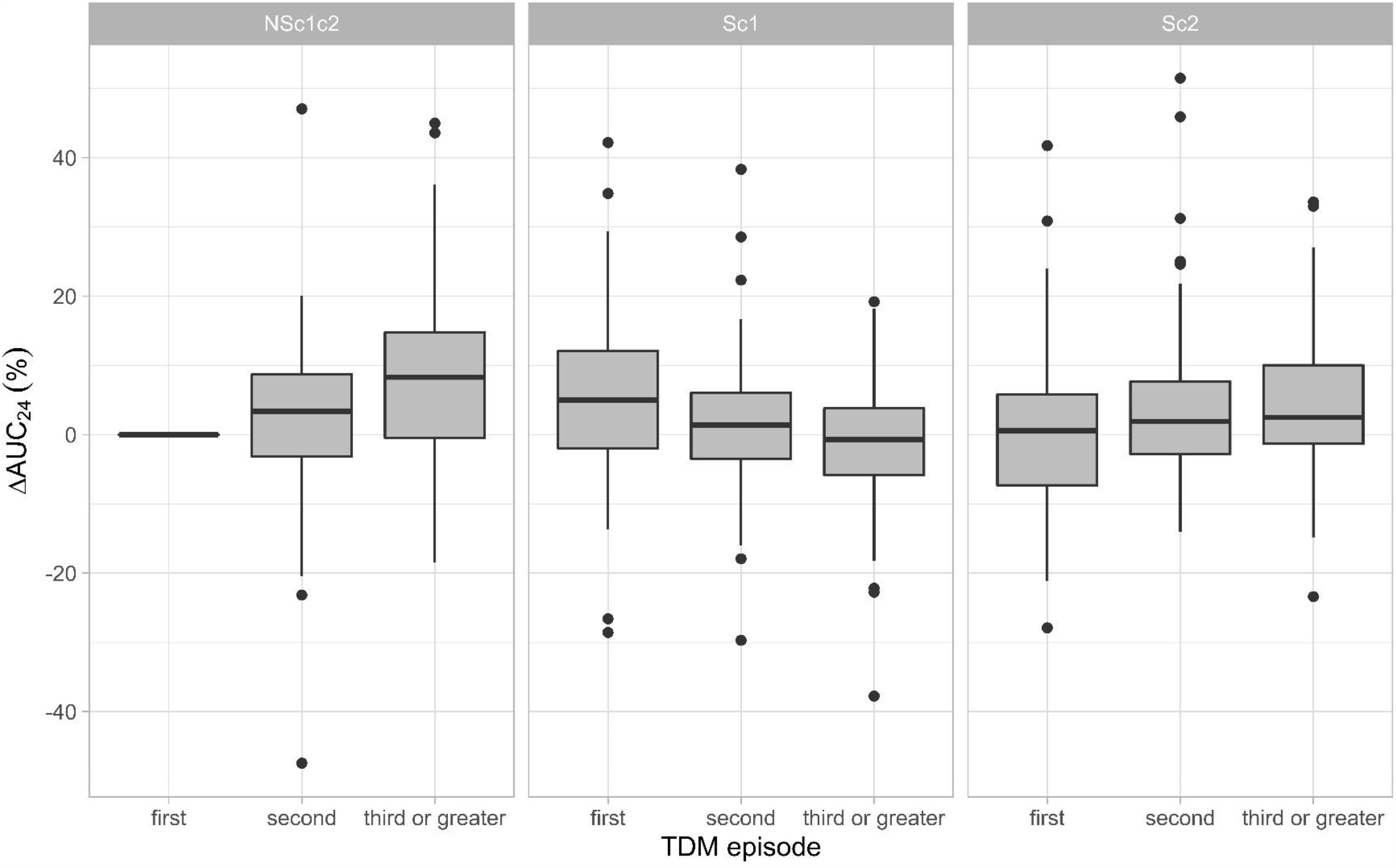
Distribution of differences in estimated AUC_24_ for full dataset (reference condition) versus selected simulation conditions according to TDM episode

## Discussion

Our study suggests that in stable adult patients with CF pulmonary exacerbations, the tobramycin AUC_24_ can be reliably estimated using Bayesian forecasting and a single post infusion concentration. In a small proportion of cases the estimates differed more significantly. The reasons for these outliers are unclear but could relate to erroneous information (e.g. time of dose or blood sample recorded incorrectly), or physiological instability. This observations illustrates the importance of attention to situations where observed concentrations are significantly different from those predicted by the model, in order to apply clinical judgement in when unexpected results occur. The graphical interface provided by modern Bayesian forecasting software allows a visual representation of this discrepancy, which may improve the identification of unusual results. Where observed concentrations deviate substantially from those predicted by the model, patients may benefit from repeat or more intensive sampling to confirm observed concentrations and to ensure an accurate estimation of their pharmacokinetic parameters.

This approach would be particularly applicable to critically unwell patients, given known physiological fluctuations in these patients where a single sample approach may be less appropriate.(16–19). Similarly, in stable patients, significantly discrepant concentrations from those expected either relative to previous estimates or predicted on the population pharmacokinetics should prompt careful consideration of a pre-analytical source of error.(20)

Although a more limited sampling approach resulted in less accurate pharmacokinetic parameter estimation relative to the reference, the effect was usually small (rRMSE <15%). Although there is no established standard for acceptable accuracy in this context, it is of comparable magnitude to accepted target ranges used in clinical practice.(12) In specific situations, e.g. those at higher risk of renal dysfunction, or unstable patients with sepsis or requiring ICU admission, a more stringent approach may be considered appropriate. In these situations, clinicians may choose to monitor the patient more closely, e.g. by continuing with two-concentration monitoring until the patient is judged stable, and then switching to a single concentration approach, e.g. to reduce phlebotomy or line access, or to facilitate ambulatory management.

Accuracy and bias of estimates were similar whether using a sample taken approximately 2 hours after the end of the infusion or approximately 6 hours after the end of infusion. These results largely agree with the results of Gao et al, who found that a single concentration taken 70 – 640 minutes yielded sufficiently accurate results.(10) For practical purposes a timed blood sample taken any time between 2-6 hours post end of infusion is likely to be sufficient for routine monitoring.

Numerous software applications for performing Bayesian forecasting are now available, however have not yet been widely-adopted, possibly owing to access to and cost of specialist software, required expertise, and the switching cost of new TDM workflows.(7,21–23) The simplicity of a single-sample Bayesian forecasting method may therefore help to offset some of these barriers for adoption. Patients with CF have generally stable tobramycin pharmacokinetics over time, and as such may be considered ideal candidates for Bayesian forecasting methods, which allow accumulation of data for an individual patient over time.(24) Patients with CF are frequently managed in CF centres and may have multiple admissions to the same institution, thus in theory clinicians managing these patients should be able to derive an accurate individualised dosing strategy for each patient. Bayesian forecasting software which allows automated population of results from an electronic medical record may therefore be of great benefit.

## 3. Conclusions

Bayesian forecasting, using a single post infusion sample between 2-6 hours post infusion and prior concentration measurements can precisely and accurately estimate tobramycin exposure. This simple approach increases the flexibility of a Bayesian TDM workflow, and may facilitate efficient outpatient monitoring for ambulatory therapy.

## Supporting information

STROBE checklist

## Data Availability

Data requests should be submitted to the corresponding author. Access to de-identified data stored on the synapse.org repository may be granted to suitably qualified researchers following review.

## Declarations

### Authors’ contributions

PD conceived the study, assisted with data collection and analysed the data and drafted the manuscript. LB collected the data and reviewed the manuscript. YT and JM performed the simulations and reviewed the manuscript. SS supported development of the TDM service and data collection, and reviewed the manuscript. SVH conceived the study and reviewed the manuscript.

